# Hard to Halt: Automation Bias in Agent-Driven Sequencing Prior Authorization Workflows

**DOI:** 10.64898/2026.06.16.26355782

**Authors:** Mengshu Nie, Wendy Chung, Jessica Waxler, Michael Lee, Chunhua Weng, Rachel Lewis, Priyanka Ahimaz, Kai Wang, Cong Liu

**Affiliations:** Boston Children’s Hospital, Department of Pediatrics, Division of Genetics and Genomics, Boston, MA; Boston Children’s Hospital, Department of Accountable Care and Clinical Integration, Boston, MA; Columbia University, Department of Biomedical Informatics, New York, NY; Columbia University Irving Medical Center, Department of Pediatrics, New York, NY; Center for Applied Genomics, Children’s Hospital of Philadelphia, Philadelphia, PA; Department of Biostatistics and Epidemiology and Department of Pediatrics, University of Pennsylvania, Philadelphia, PA; Boston Children’s Hospital, Children’s Hospital Informatics Program, Boston, MA; Harvard Medical School, Department of Pediatrics, Boston, MA

**Keywords:** Prior authorization, Large language models, Browser agents, Agent performance Evaluation

## Abstract

**Purpose:** Prior authorization (PA) for exome or genome sequencing (ES/GS) is a time-consuming process that impedes timely rare disease diagnosis. Large language model (LLM)-based browser agents offer potential for automating these workflows, but their clinical reliability remains uncharacterized.

**Methods:** We built a sandbox with a simulated ES/GS payer portal and a synthetic electronic health record (EHR) of 836 patient profiles, including compliant profiles and deficient profiles with designed issues. Gemini 3 Pro, Gemini 3 Flash, and Claude Opus 4.5 were evaluated on task completion, form accuracy, and appropriate withholding. Two replication experiments were run on 100 profiles to assess run-to-run variability of agent performance.

**Results:** Gemini 3 Pro (95.5%) and Claude Opus 4.5 (93.7%) completed far more tasks than Gemini 3 Flash (56.1%), consistently across runs. In the first run, all models rarely withheld deficient profiles for the correct reason (Gemini 3 Pro 0%, Gemini 3 Flash 17.3%, Claude Opus 4.5 5.2%). Replication revealed substantial variability: Gemini 3 Pro’s correct withholding rose to 28.8–36.8%, and only 18.0–48.0% of tasks yielded identical outcomes across all three runs. Without agentic scaffolding, Gemini 3 Pro flagged designed issues in 91.3% of deficient profiles but also halted on all compliant profiles.

**Conclusion:** Current LLM-based browser agents show a systematic bias toward form submission. A modular, multi-agent architecture with human supervision may be warranted for safe clinical deployment.

## Introduction

Prior authorization (PA) is an insurer-initiated process to verify medical necessity and coverage eligibility before approving prescribed procedures, services, or medications requested by healthcare providers.^1^ It has contributed significantly to healthcare administrative costs in the United States, estimated to account for $35 billion annually.^2^ These costs are driven in part by the high frequency of administrative errors in PA submissions, including missing documentation, coding mistakes, and eligibility mismatches, which necessitate resubmission, appeal, and manual review.^3^ Across *Affordable Care Act* (ACA) Marketplace plans, such administrative reasons accounted for 25% of all in-network claim denials in 2024, substantially exceeding denials based on medical necessity (5%).^4^ These failures are often traceable to inaccuracies propagated from the Electronic Health Records (EHR), where incomplete or incorrect data are incorporated into PA submissions without adequate verification.^5^ Beyond the monetary costs, the PA process is also time-consuming. An American Medical Association survey in 2024 showed that each physician and their staff spend an average of 13 hours on PA forms per week.^6^ This burden is particularly acute for clinical exome and genome sequencing (ES/GS), which have transformed rare disease diagnosis by increasing diagnostic yield and reducing cost per diagnosis.^7,8^ Obtaining PA approval for these genetic tests, especially in outpatient contexts, is a significant bottleneck: approximately 20% of 4,535 PA requests for genetic testing at two children’s hospitals in Texas were denied, with ES having the second-highest denial rate among all genetic tests.^9^ Denials also carry clinical consequences: in a pediatric genomic research cohort of 801 patients, 18.3% had a PA denial on record; among these patients, 53.7% had at least one diagnostic or candidate finding, 10.9% had a clinically diagnostic finding, and half of those with diagnostic results had immediate changes in medical management.^10^

The follow-up appeal process after an initial denial can introduce weeks of additional delay, which further impedes timely diagnosis and treatment for patients.^11^

Currently, most PA submissions still rely on manual web portals or fax, which contribute to transcription errors and inefficiency. According to the 2024 CAQH Index, only 35% of medical PA transactions were fully electronic, with the remaining 65% split between web portals (43%) and fully manual channels including phone, fax, mail, and email (22%).^12^ Large language model (LLM)-based agents offer a potential solution to these administrative challenges for the genetic counselors, physicians, and medical assistants who currently bear the operational burden of PA submission in genetics practices. Unlike static conversational systems, agentic LLMs execute multi-step tasks by iteratively reasoning, planning, and acting through external tools. The ReAct framework formalizes this approach by coupling stepwise reasoning with discrete actions, enabling interaction with application programming interfaces (APIs), databases, and web interfaces.^13^ Browser-based agents operationalize this paradigm for web workflows by navigating pages, parsing web representations and selectively incorporating visual context via vision-language models when needed.^14^ For PA submission, this enables end-to-end automation: logging into payer portals, navigating form fields, filling in structured EHR data (demographics, codes, insurance details), and submitting requests with confirmation capture. This closed-loop interaction aligns naturally with the structured, form-based design of PA systems, making ES/GS PA a suitable target for agentic automation.

The evaluation of LLM agents in clinical and healthcare settings has become an active area of investigation. General-purpose agent benchmarks such as WebArena^15^ established methodological foundations for evaluating web-navigating agents on realistic tasks, and medical-domain benchmarks including MedAgentBench^16^ have extended these methods to EHR environments. More recently, HealthAdminBench^17^ introduced a broad evaluation framework specifically for computer-use agents on healthcare administrative workflows including prior authorization, appeals and denials management, and durable medical equipment order processing. The authors reported 36.3% end-to-end task success for the best-performing agent, concluding that current frontier agents fall well short of the reliability required for autonomous deployment. Our study complements this body of work: rather than benchmarking across multiple administrative tasks, we focus on a specific mode of failure: whether agents correctly identify and withhold submission on PA forms when errors or issues are present in patient information. Because agentic systems, unlike conversational tools, can execute consequential and irreversible actions, such as modifying EHR-linked records or disclosing protected health information, their behavior must be rigorously examined in a controlled environment before clinical deployment. In this study, we investigate the behavior of LLM-based browser agents in a controlled sandbox environment simulating the ES/GS PA workflow, characterize their task completion performance, identify systematic failure patterns, and establish a quantitative foundation to guide future clinical implementation.

## Materials and Methods

### Sandbox Environment & Patient Data

We developed a sandbox environment comprising two primary components (***Figure 1(a)***. First, a payer web portal with a PA form was constructed to emulate real-world payer forms, requiring structured inputs including patient demographics, insurance subscriber information, provider and laboratory details, and relevant clinical data. Current Procedural Terminology (CPT) codes are retrieved through a built-in search function within the form rather than from the patient record. We then implemented a searchable patient database that simulates an EHR environment from the provider’s side. Note that the patient record is not intended to replicate a complete chart from a real-world EHR system; instead, each patient profile is consolidated into a single structured information card which provides text-based information needed to complete the PA form. The sandbox is publicly available at https://wes-wgs-pa-app-u2c8s.ondigitalocean.app/login.

**Figure 1.**
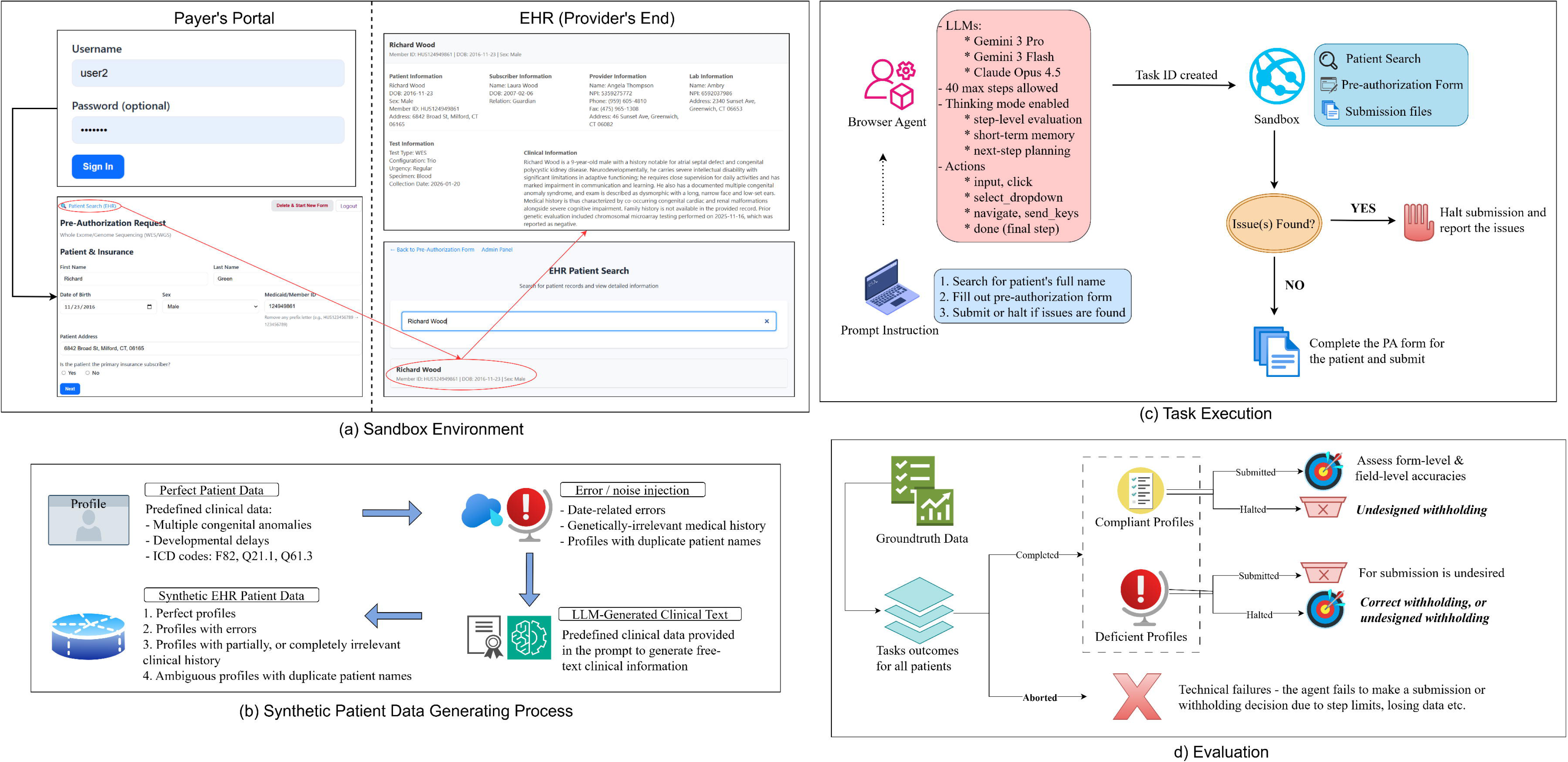
Sandbox & Study Overview. (a) The sandbox interface consists of a user login page, a PA form, and a patient search function that directs to the synthetic EHR database. The payer portal requires authentication prior to form access. The EHR interface supports patient retrieval via name-based search, and each patient record contains structured data fields (demographics, subscriber, provider, laboratory, and test details) alongside a free-text clinical summary. (b) Patient profile generation process. All profiles were generated from predefined clinical condition templates and ICD codes, and subsequently augmented with LLM-generated free-text clinical narratives conditioned on the structured data. (c) A browser-based agent was initialized with a standardized prompt, and a unique task ID was automatically assigned at the time of task creation. Each task was executed in an independent session within the controlled sandbox environment per patient profile and per model, with no shared state or memory across runs. A maximum of 40 browser interaction steps were enforced per task, and the thinking mode was enabled to produce explicit intermediate reasoning traces, including step-level evaluation, short-term memory, and next-action planning. (d) Task outcomes for all patient profiles are evaluated against ground-truth data. For compliant profiles, submitted forms are assessed for form-level and field-level accuracy; halted submissions are flagged as over-refusal. For deficient profiles, submission is considered undesired, while halted submissions can be either correct withholding or error misattribution depending on whether the identified issue matches the ground-truth injected error. Tasks that terminate without producing either a submission or withholding decision, such as due to step limit exhaustion, data loss, or other technical failures, are classified as *aborted*.

A synthetic patient cohort was then constructed via a multi-stage data generation pipeline as visualized in ***Figure 1(b)***. To emulate the variability observed in real-world clinical data and evaluate agent performance across different data contexts, we partitioned the patient profiles into four groups (Table 1). Profiles in categories 2, 3, and 4 were classified as deficient because submission should be halted due to injected errors. Homonymous profiles (category 4) contain no errors themselves; they are deficient because the agent receives only a patient name, which makes patient search ambiguous. Details of the data generation pipeline and data fields are provided in the Supplementary Material (Patient Data Generation; Supplementary Table S1). The PA form and the composition of patient records were reviewed by a clinical genetic counselor to ensure that the information available in the sandbox was at least consistent with what would inform a real PA decision.

**Table 1.** Patient Profile Categories.

| Profile Category | Subcategory | Label | Description | Example |
|---|---|---|---|---|
| Compliant profiles | Exclusive genetic relevancy (n = 105) | 1a | Profiles with no designed errors and clinical description are entirely related to genetic conditions | N/A |
|  | Mixed genetic relevancy (n = 107) | 1b | The clinical information is primarily related to genetic disorders / phenotypes but also mixed with unrelated secondary medical issues (e.g., concussion, physical injuries) | The clinical information describes the patient’s phenotype of congenital malformation syndrome and intellectual disability, as well as physical injuries and burning as secondary medical history |
| Error-injected profiles | Subscriber age error (n = 107) | 2a | Subscriber date-of-birth error:<br>the subscriber is too young to be the patient’s parent / legal guardian based on age gap | Patient date-of-birth: 20-01-2012<br>Subscriber date-of-birth: 08-01-2003 |
|  | Chronological error (n = 105) | 2b | Chronological errors: the prior test date provided in the clinical record is after the specimen collection date for ES/GS | Prior test date: 02-03-2026<br>Specimen collection date: 15-02-2026 |
|  | Missing date (n = 106) | 2c | Missing required date information | Specimen collection date (required in the PA form) is intentionally labelled as *“N/A”* in the patient record |
| Irrelevant profiles | N/A (n = 106) | 3 | The clinical information exclusively describes genetically irrelevant medical history | The clinical information only describes the patient’s past medical history about poisoning and burning |
| Homonymous profiles | N/A (n = 200) | 4 | Profiles occur in pairs that share an identical patient name | N/A |

ES, exome sequencing; GS, genome sequencing; PA, prior authorization. Dates are shown in DD-MM-YYYY format.

### Agent Setup and Task Execution

PA submissions were automated using *Browser Use Cloud* (***Figure 1(c)***), the hosted service of an open-source agentic framework which enables LLMs to orchestrate browser-based workflows such as webpage navigation and information retrieval. Three models were evaluated: Gemini 3 Pro, Gemini 3 Flash, and Claude Opus 4.5. Gemini 3 Pro and Claude Opus 4.5 were selected as representative frontier models because they completed trajectories more reliably than other models available through Browser Use in preliminary testing, including OpenAI’s GPT-4.1 and o3. Gemini 3 Flash was included as a lighter-tier comparison within the same model family. For each patient profile, the agent was initialized with a baseline prompt (**Supplementary Prompt *2***) without explicit guidance regarding potential error types or fields requiring scrutiny. Each task was limited to 40 browser interaction steps, with thinking mode enabled. In addition to the main experiment, we conducted two replication runs on a stratified subset of 100 profiles and three ablation studies (Table 2; details in Results). Model identifiers, Browser Use API versions, and run dates for all experiments are listed in *Supplementary Table S4*.

**Table 2.**
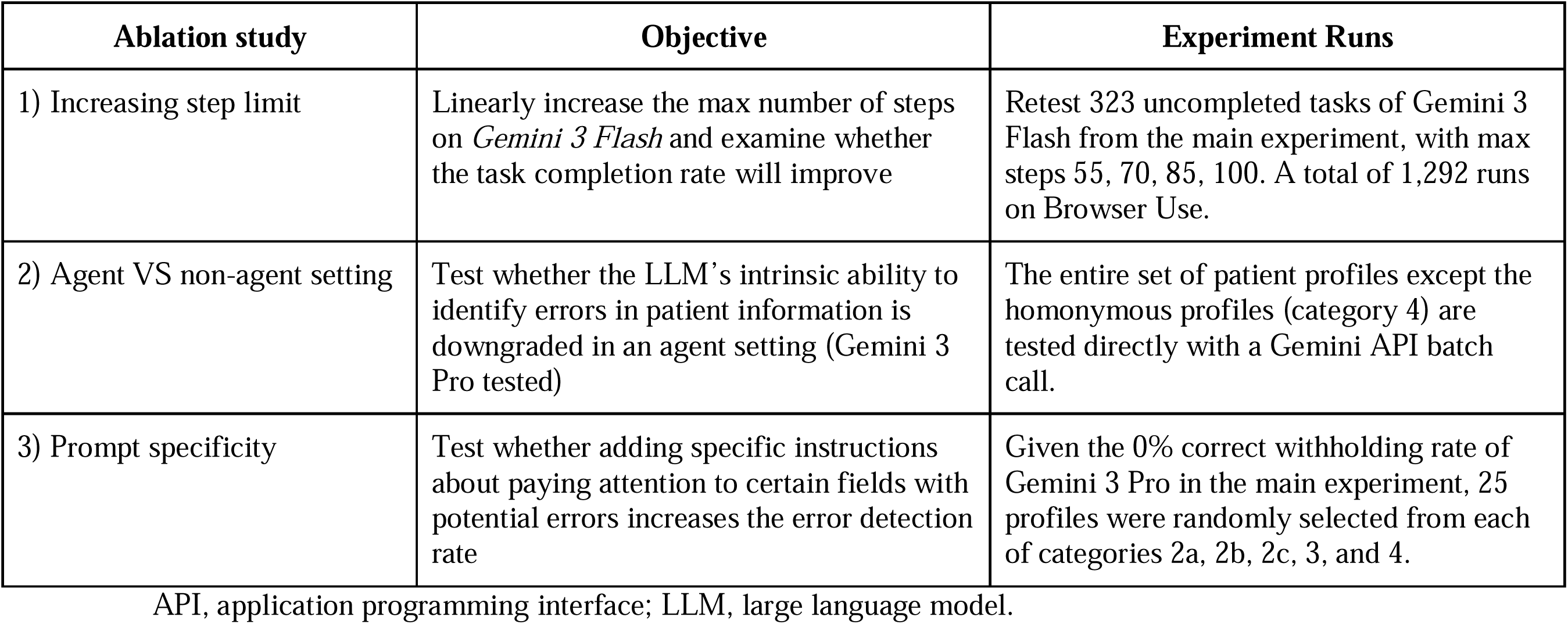
Ablation Study Summary.

| **Ablation study** | **Objective** | **Experiment Runs** |
|---|---|---|
| 1) Increasing step limit | Linearly increase the max number of steps on *Gemini 3 Flash* and examine whether the task completion rate will improve | Retest 323 uncompleted tasks of Gemini 3 Flash from the main experiment, with max steps 55, 70, 85, 100. A total of 1,292 runs on Browser Use. |
| 2) Agent VS non-agent setting | Test whether the LLM’s intrinsic ability to identify errors in patient information is downgraded in an agent setting (Gemini 3 Pro tested) | The entire set of patient profiles except the homonymous profiles (category 4) are tested directly with a Gemini API batch call. |
| 3) Prompt specificity | Test whether adding specific instructions about paying attention to certain fields with potential errors increases the error detection rate | Given the 0% correct withholding rate of Gemini 3 Pro in the main experiment, 25 profiles were randomly selected from each of categories 2a, 2b, 2c, 3, and 4. |

API, application programming interface; LLM, large language model.

## Evaluation

***Figure 1(d)*** illustrates the evaluation framework used to assess agent performance across profile types. We first classified the agents’ task execution outcomes into one of the following categories:

1. *Aborted*: the agent ended without submitting the form or giving a reason for withholding, because of a technical failure such as reaching the 40-step limit or losing form data after a browser refresh.
2. *Submitted*: the agent completed and submitted the PA form. This is the correct outcome for compliant profiles and an error for deficient profiles.
3. *Correct withholding*: the agent halted on a deficient profile and reported its designed issue. This is the desired outcome for deficient profiles.
4. *Over-refusal*: the agent halted on a compliant profile for an unsupported reason, such as claiming that information present in the record was missing or failing to infer International Classification of Diseases (ICD) codes or clinical rationale that could be derived from the record.
5. *Error misattribution*: the agent halted on a deficient profile but cited an issue other than the designed one.

Example outputs for each outcome are provided in *Supplementary Table S2*. Task-level metadata were retrieved via the Browser Use API, and submitted cases were identified by cross-referencing the full set of task IDs against the submission files stored in the sandbox.

For tasks without a recorded submission, GPT-5.2 was then applied to the agent’s final output messages to categorize outcomes into one of the four outcome labels above (**Supplementary Prompt *3***). The GPT-classified labels showed almost perfect agreement with blinded human annotation on a stratified sample of 174 outputs (Cohen’s κ = 0.89; Supplementary section, *Human VS GPT-5.2 Classification* & Supplementary Table S3). We then evaluated form-filling accuracy on submitted compliant profiles by comparing each agent-filled value against its ground-truth entry. Fields were grouped as deterministic, whose values can be retrieved directly from the patient record (e.g., demographics, provider, laboratory, and test details), or interpretative, which require clinical reasoning or inference from narrative text (e.g., rationale for testing and prior testing history). Uncertainty in outcome rates and accuracies was estimated by nonparametric bootstrap resampling with 1,000 iterations.

To contextualize agent performance against human judgment on the same task, we recruited a genetic counselor to independently complete PA submissions through the same sandbox interface, following the same halting instructions given to the agents and blinded to profile categories and issue types. Since manual completion of the full cohort was not feasible, we only drew a stratified subset of 30 profiles (*Supplementary Material, Human Baseline*).

## Results

### Task Execution Outcomes

***Figure 2(a)*** shows task execution outcomes from the first full-cohort run (main experiment). The frontier models exhibited substantially fewer technical failures, resulting in much higher overall task completion rates (95.5% for Gemini 3 Pro and 93.7% for Claude Opus 4.5) compared to only 56.1% for Gemini 3 Flash. Among completed tasks, both Gemini models achieved high submission rates for compliant profiles (99.5% for Gemini 3 Pro, 91.5% for Gemini 3 Flash) whereas Claude Opus 4.5 over-refused 32.1% of compliant profiles, for example by stating that provider information was missing when it was present. For deficient profiles, all three models largely failed to identify injected issues and appropriately halt execution. Correct withholding rates in this run were 0% for Gemini 3 Pro, 5.2% for Claude Opus 4.5, and 17.3% for Gemini 3 Flash. Although Claude Opus 4.5 withheld the highest proportion of deficient profiles (37.4%), it mostly halted for misattributed reasons (32.2%) rather than the designed issues. Figure 2(b) breaks down outcomes on deficient profiles by issue type. Both Gemini 3 Flash and Claude Opus 4.5 correctly withheld profiles with a missing collection date (Gemini 3 Flash 32.0%, Claude Opus 4.5 10.0%) or irrelevant clinical history only (Gemini 3 Flash 39.0%, Claude Opus 4.5 17.0%).

**Figure 2.**
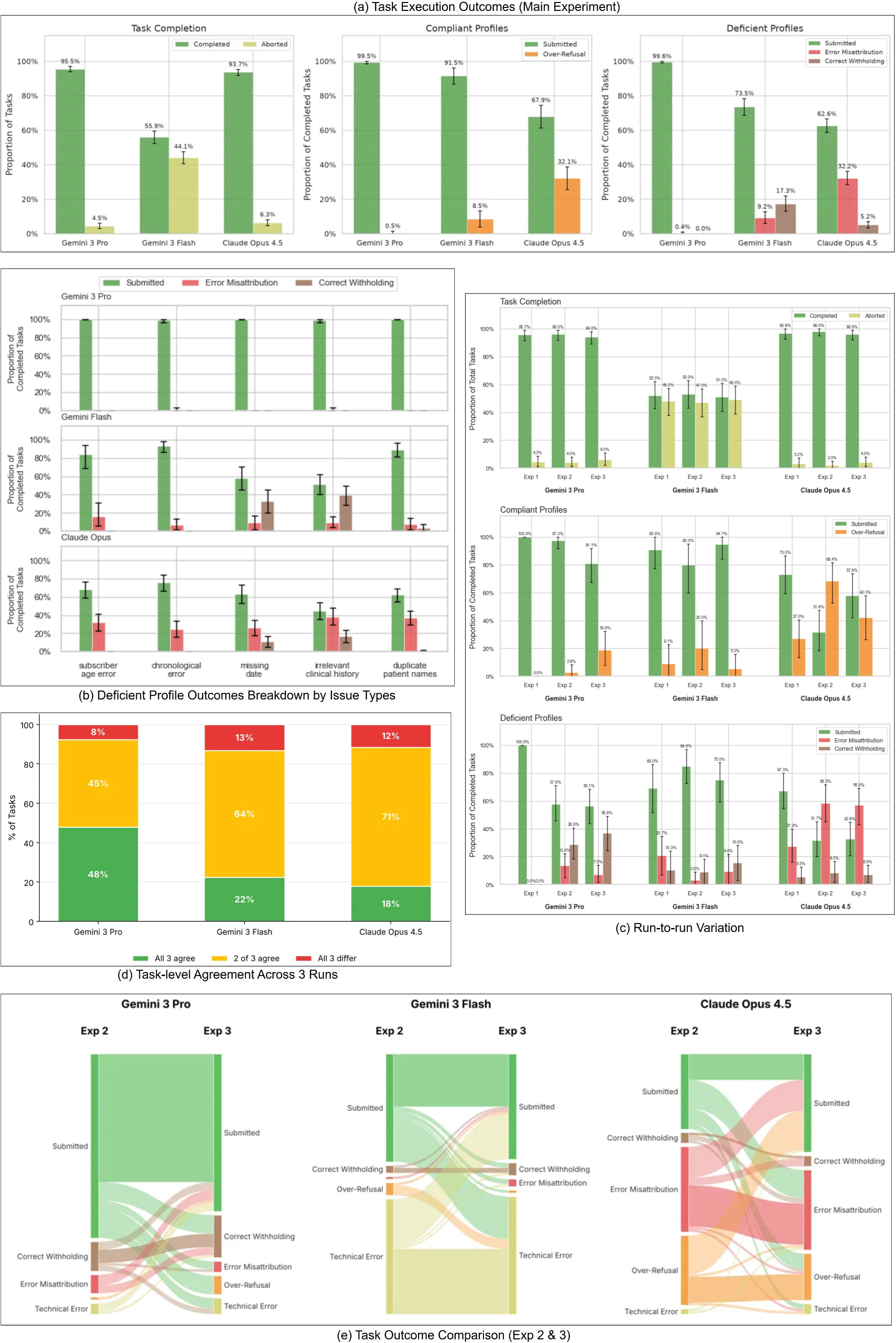
Main Experimental Results. (a) Left: Proportions of completed / aborted tasks. Middle & Right: Proportions of other task outcomes conditioned on the completed tasks across compliant and deficient profiles, with error bars indicating 95% bootstrap confidence intervals (1,000 iterations). (b) Task outcome breakdown by issue types for deficient profiles. (c) Task outcome comparison among three runs on the same subset of patient profiles. Exp 1 indicates the original experiment, Exp 2 and 3 are the subsequent replication runs. (d) Per-task outcome consistency across the three runs. Each bar shows the proportion of profiles on which all three independent runs produced the same outcome category (green), two of three runs agreed (yellow), or all three runs differed (red), for each of the three models. (e) Per-task outcome transitions between replication runs (Exp 2 & Exp 3). Alluvial diagrams show how each profile’s outcome changed from Exp 2 to Exp 3. Bands that stay straight indicate the same outcome in both runs, crossing bands indicate a change, and band width is proportional to the number of profiles.

### Run-to-run Variability in Task Outcomes

To examine run-to-run variation, we conducted two additional replication runs on a stratified subset of 100 profiles using the same Browser Use harness version and API model identifiers (Figure 2(c)). Task completion rates were relatively stable across the three runs (Gemini 3 Pro 94.0–96.7%, Gemini 3 Flash 51.0–53.0%, Claude Opus 4.5 96.0–98.0%), but outcome distributions among completed tasks varied. The qualitative pattern of our primary findings still holds across all three runs: Claude Opus 4.5 exhibits the highest error misattribution and over-refusal rates, and all three models predominantly failed to withhold submission for the correct designed issues on deficient profiles. However, the specific proportions differed between runs. The most pronounced shift is observed for Gemini 3 Pro on deficient profiles, where the original experimental run (Exp 1) showed 100% submission with 0% correct withholding while both later replicate runs showed substantially higher correct withholding rates (28.8% and 36.8%). Notably, the replicate runs were conducted 7–8 months after the main experiment; this shift may therefore reflect provider-side updates to the underlying model, residual stochasticity in model outputs on identical inputs, or both. Similar temporal drift and run-to-run nondeterminism have also been reported for LLMs applied to genetic variant assessment.^18^ *Figure 2(d)* shows variability at the level of individual tasks: fewer than half of tasks produced identical outcomes across all three runs (Gemini 3 Pro 48.0%, Gemini 3 Flash 22.0%, Claude Opus 4.5 18.0%). Even Exp 2 and Exp 3, conducted together on the same model state, showed frequent task-level flipping (*Figure 2(e)*), with pairwise agreement of only 58.0% (Gemini 3 Pro), 50.0% (Gemini 3 Flash), and 39.0% (Claude Opus 4.5). Thus, agent outcomes on fixed inputs can be substantially non-deterministic, and stable aggregate percentages do not necessarily imply stable task-level outcomes.

### Error Analysis

To understand why agents halted for reasons other than the designed issues, we manually reviewed all over-refusal and error misattribution cases from the *main experiment* for Claude Opus 4.5 (n = 244) and Gemini 3 Flash (n = 37). Gemini 3 Pro produced only 3 such cases and was not analyzed further. For Claude Opus 4.5, 93.4% of the cited issues were hallucinations (e.g., claiming that the subscriber date of birth or provider information was missing from the patient record) and 6.6% were inference failures (e.g., reporting ICD-10 codes or clinical rationale as absent when they could be inferred from the clinical narrative), whereas for Gemini 3 Flash, inference failures accounted for 56.8% and hallucinations for 43.2%.

To identify the mechanisms behind these outcomes, we further examined 24 full agent trajectories from the main experiment and replication runs (two per model and non-submitted outcome category), from which a few recurring patterns emerged. First, the hallucinations described above appeared to arise from how agents managed information retrieval rather than from fabrication. In 5 of 6 error misattribution cases and 3 of 6 over-refusal cases, the agents halted because a field was “missing from the EHR,” even though it was present in the record.

In these trajectories, the field had been omitted when the agent first transcribed the patient record into its working memory or a local notes file; the agent subsequently treated this transcribed state as the only source of data and did not re-consult the EHR. Second, in 11 of 24 trajectories (all 8 Gemini 3 Flash runs, 2 Gemini 3 Pro, and 1 Claude Opus 4.5), the agent fragmented data retrieval into repeated round trips between the patient record and the form, reopening the record once per form section rather than extracting all information in a single pass. This pattern consumed a substantial share of the 40-step budget and likely contributed to the high aborted rate observed for Gemini 3 Flash. Third, in the 3 trajectories involving genetically irrelevant profiles, the agent successfully extracted the clinical narrative but proceeded to form filling without evaluating whether it supported ES/GS testing, suggesting that relevant content in context was not used to evaluate medical necessity. Given the small sample, these observations characterize recurrent patterns only qualitatively and do not support cohort-level frequency estimates.

### Submitted Form Accuracy

***Figure 3(a)*** shows the bootstrapped distribution of form-level accuracy on submitted compliant profiles; all three models exceeded 90% (Gemini 3 Pro 94.4%, Claude Opus 4.5 93.7%, Gemini 3 Flash 91.1%). ***Figure 3(b)*** presents accuracy for each required field. All models achieved near-ceiling accuracy in most deterministic fields but substantially lower accuracy on a few others. For example, the frontier models occasionally inferred a parental subscriber relationship when the record indicated “other” or “guardian,” whereas Gemini 3 Flash sometimes left subscriber fields blank despite explicit information. Sporadic hallucinations were observed for all three models (7.6% for Claude Opus 4.5, 5.7% for Gemini 3 Pro, and 4.2% for Gemini 3 Flash, computed over submitted tasks), with address-related fields populated with entirely fabricated values. Accuracy was more variable for fields requiring clinical interpretation: rationale-for-testing checkboxes remained relatively accurate (80.0–100.0%), whereas prior testing history fields, which require extraction from narrative text, showed the lowest accuracy (50.0–80.0%).

**Figure 3.**
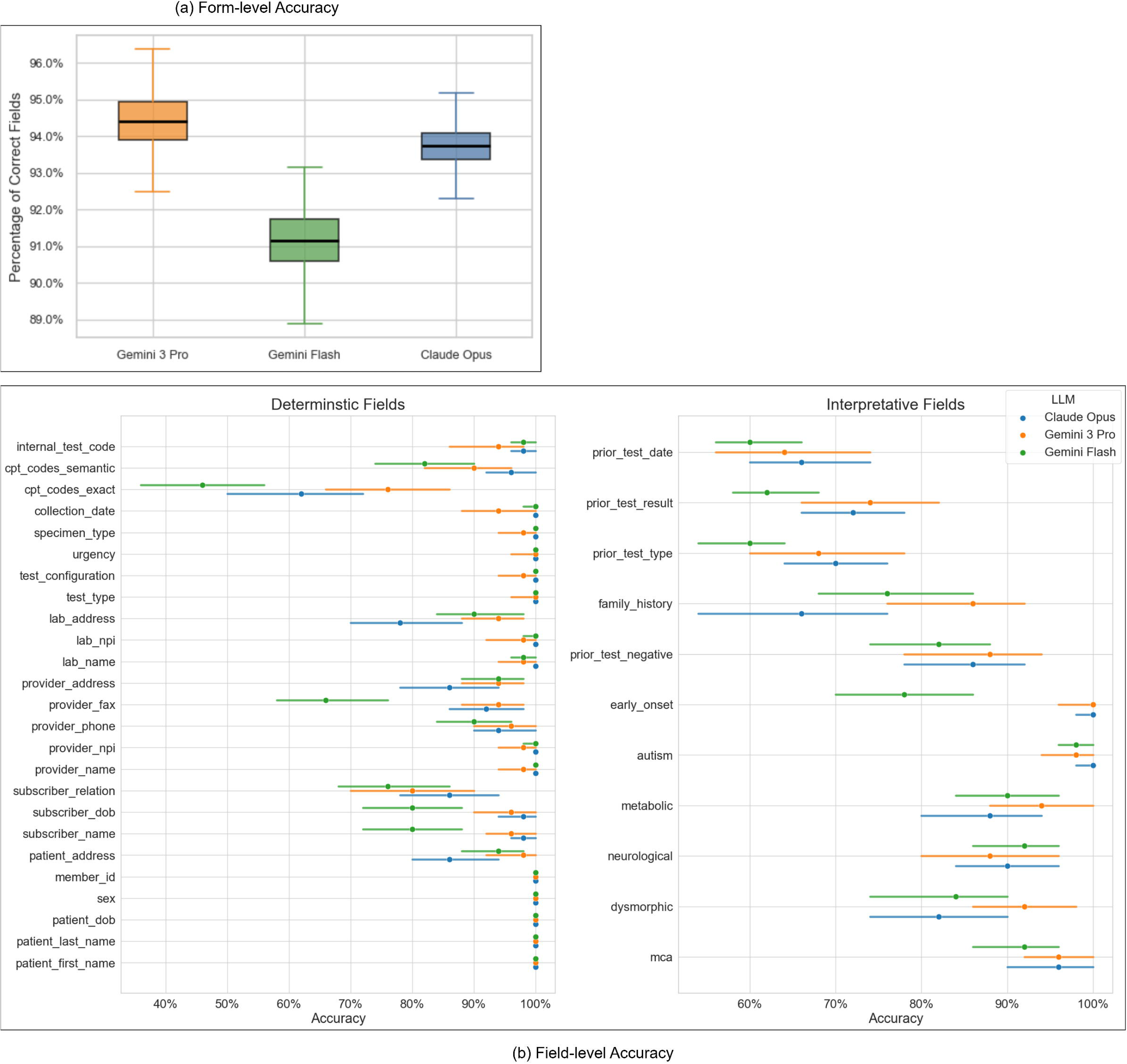
Form-filling Accuracy. (a) Proportions of correctly completed fields per submission among successfully submitted compliant profiles. (b) Field-level accuracy (median) across all required form fields with 95% bootstrap confidence intervals (1,000 iterations), i.e., the proportion of submitted forms in which each field was completed correctly.

### Human Baseline

In the human baseline experiment, the genetic counselor’s median completion time was 8.82 minutes per profile, compared with 4.47 min for Gemini 3 Pro, 5.40 min for Gemini 3 Flash, and 8.14 min for Claude Opus 4.5 (computed over completed tasks). The counselor submitted all compliant profiles and readily identified all profiles with clinically irrelevant indications for ES/GS testing (profile category 3) but did not identify any of the other issue types. On profiles with a missing specimen collection date, the counselor entered the current date and proceeded with submission rather than reporting the missing field. This mirrors the agents’ behavior on the same profile type: in cases where agents submitted these profiles, the missing field was also filled with the date when the form was accessed.

### Ablation Studies

Three ablation studies were further conducted to examine the impact of step budget, the agentic setting, and prompt specificity on agent performance (Table 2).

### Ablation 1: increasing step limit

Gemini 3 Flash demonstrated the highest rate of technical failures in the main experiment, likely because of repeated round trips between the patient record and the PA form (see Error Analysis). We therefore re-ran Gemini 3 Flash on the 323 of 735 tasks that had not completed at 40 steps, with step limits of 55, 70, 85, and 100. At 55 steps, 89.2% of these previously uncompleted tasks completed (*Figure 4(a)*); further increases yielded diminishing gains, with completion saturating above 99% at 85 and 100 steps. Form-level accuracy (Figure 4(b)) was highest at 55 steps (median 95.8%) and declined as the budget was relaxed, suggesting that looser budgets, while allowing more tasks to finish, may introduce noise through redundant re-interactions with already-completed fields.

**Figure 4.**
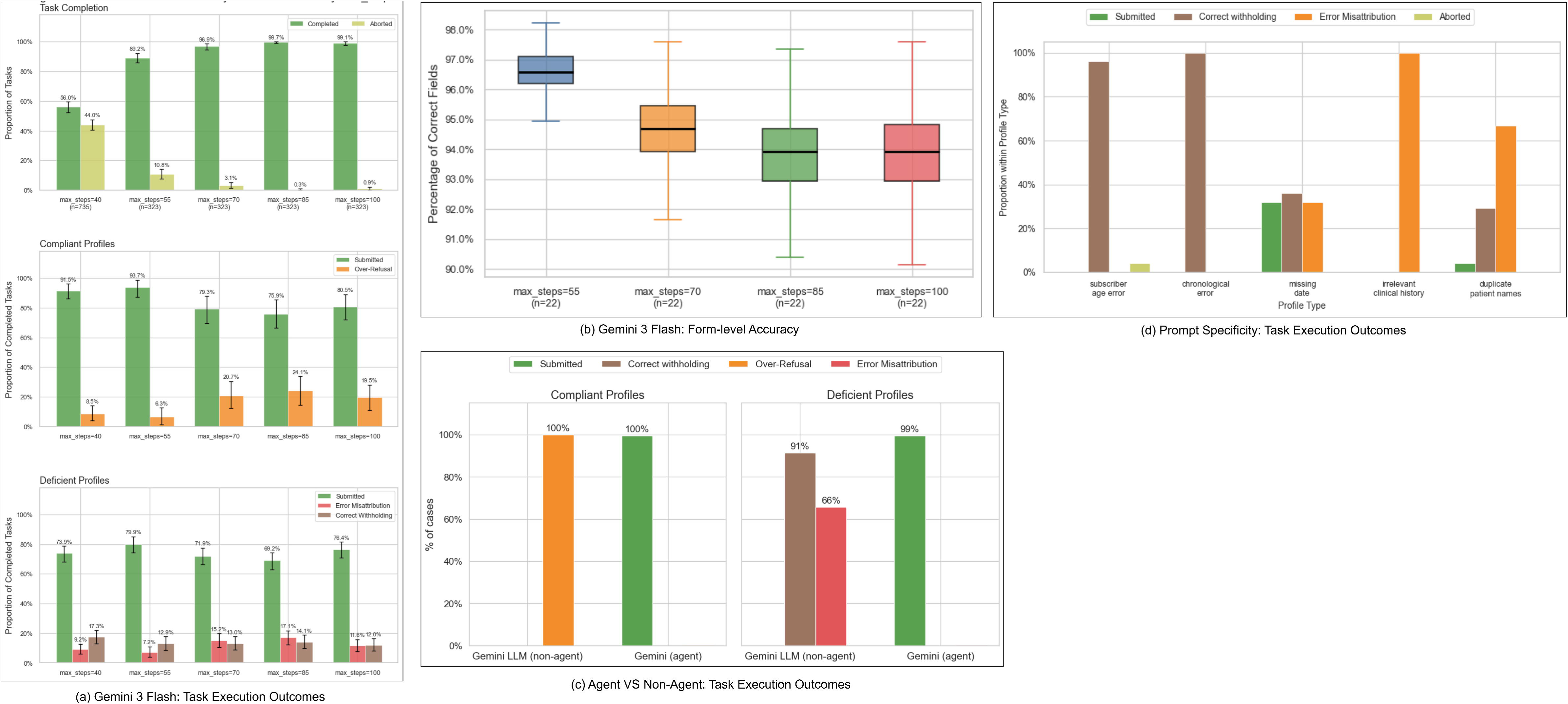
Ablation studies. Ablation Study 1: Given the high rate of technical failures observed for Gemini 3 Flash under a 40-step constraint in the main experiment, we systematically increased the maximum allowed steps in this ablation study. The 40-step bar reflects all 735 tasks; the 55-, 70-, 85-, and 100-step bars reflect only the 323 tasks that failed to complete at 40 steps. (a) Left: task completion rate (bootstrapped median with 95% confidence intervals) as a function of step limit. Right: distribution of task outcomes among completed tasks. (b) Proportions of correctly completed fields per submission among successfully submitted compliant profiles across different step limits. Ablation study 2: (c) Comparison between agent-based and non-agent (chat-only) settings. In the non-agent condition, a similar prompt with patient profiles was provided to Gemini 3 Pro in a chat API to elicit submission or withholding decisions along with identified issues. Task outcomes are compared across settings. Technical errors are not applicable in the non-agent condition due to the absence of browser execution. For deficient profiles, the model may identify both injected (ground-truth) and undesigned issues within a single instance; therefore, proportions do not sum to 100%. Ablation study 3: (d) Agent performance on 125 random deficient profiles (25 profiles from each of categories 2a, 2b, 2c, 3, and 4) with enhanced prompt instructions (e.g., explicit emphasis on reviewing date-related fields). The panel reports the distribution of task outcomes across profile categories.

### Ablation 2: agent VS non-agent setting

To disentangle the contribution of agentic scaffolding from the underlying LLM’s intrinsic behavior on the same inputs, we compared outcomes between the full agent setting and a non-agent baseline across all profiles except homonymous profiles (category 4), using Gemini 3 Pro as a representative model. For the non-agent setting, we provided the complete patient profile in dictionary format directly in the prompt and asked the model to make a submission or withholding decision (**Supplementary *Prompt 4***) without any additional workflow scaffolding. A preliminary manual review of a subset of responses revealed two patterns not observed in the agentic setting: 1) the model flagged inherent characteristics of the synthetic data in both compliant and deficient profiles, such as provider National Provider Identifier (NPI) numbers deviating from real-world formatting conventions and addresses not matching known geographic entries; 2) the model often reported both designed and non-designed issues for the same deficient profile, so that correct withholding and error misattribution applied simultaneously. We therefore used a modified classifier prompt that allowed both labels (*Supplementary Prompt 5*). *Figure 4(c)* shows that the model withheld 100.0% of the profiles regardless of the compliance status and correctly identified the designed issue in 91.3% of deficient profiles; 62.3% of these outputs with correct identifications also cited alongside at least one other non-designed issue. Out of all deficient profiles, synthetic-data related artifacts were cited in 29.7% of the total responses (25.9% of compliant and 31.6% of deficient profiles). Excluding these artifacts, the model also cited at least one issue other than the designed error in all the compliant profile responses and 59.7% of the deficient profile responses.

### Ablation 3: prompt specificity

Given the low correct withholding rates observed for profiles with injected issues or errors, we tested whether targeted prompt augmentation could improve error detection. Specifically, the phrase *“carefully review date-related information in the patient record”* was appended to the standard prompt used for the main experiment (**Supplementary *Prompt 6***) and evaluated on 125 *randomly* selected deficient profiles (25 per category) using Gemini 3 Pro as a representative model, which had shown 0% correct withholding in the main experiment.

Since this ablation was conducted within the same time window as the main experiment (*Supplementary Table S4*), the main-experiment outcomes on the same profiles serve as a concurrent baseline. As shown in *Figure 4(d)*, this prompt modification produced substantial improvements in correct withholding for date-sensitive profiles: profiles with subscriber age error improved to 96.0% correct withholding, and profiles with chronological errors reached 100.0%. However, the benefit was not uniform: profiles with missing specimen collection date, a required field in the PA portal, reached only 36.0% correct withholding, and the improvement did not generalize to non-date-specific issues (0% correct withholding for profiles with irrelevant clinical history and 29.2% for profiles with duplicate patient names). Manual review of agent outputs on the clinically irrelevant profiles clarified the mechanism: agents consistently halted by citing the specimen collection date as being in the past, reasoning that a pre-authorization cannot be submitted for a procedure that has already taken place. This is a misconception about the PA workflow: the service under authorization is the sequencing test itself, and specimen collection prior to authorization is routine in practice.

## Discussion

In this study, we developed a sandbox and evaluation framework for LLM-based browser agents performing ES/GS PA submission, focusing on whether agents halt properly when patient information contains errors. This capability is critical given the prevalence of clinical documentation errors,^19,20^ yet it remains understudied in existing agent benchmarks^15,21–23^. Across the three models evaluated, agents rarely withheld submission for the correct reason, reflecting a systematic bias toward form submission that poses operational and compliance risks in PA workflows. The human baseline suggests that the gap between agents and a human reviewer may be narrower for administrative errors, which the genetic counselor also largely missed, than for assessments of clinical appropriateness. Because the baseline comprised a single reviewer on 30 profiles, a larger multi-reviewer study would be needed to establish a formal human-performance benchmark.

The sandbox used in this study simplifies the real PA workflow and EHR environment, as reproducing a production EHR was not feasible within the scope of this study. This simplification can, however, be viewed as a lower bound on task difficulty that sets a minimum standard for agent performance: if an agent cannot reliably identify issues when the information is presented in a structured and searchable format, it is unlikely to perform better in a real-world setting. The realistic clinical deployment of an agentic PA pipeline would therefore look substantially different from the sandbox. Patient information should be retrieved programmatically from the institution’s EHR through a standard interface such as Health Level Seven (HL7) Fast Healthcare Interoperability Resources (FHIR), which exposes structured resources of each individual patient without having to navigate EHR user interfaces. The agent would then interact with the payer portal only for form filling, a step naturally suited to browser-based automation.

Our agent experiment results carry several important implications. First, Ablation 2 suggests that, outside an agentic workflow, the same model scrutinizes inputs broadly and flags anomalies indiscriminately, and that this scrutiny is suppressed to some extent once the model is embedded in a workflow focused on task execution. The error analysis points in the same direction: in clinically irrelevant profiles, agents extracted the clinical narrative but did not evaluate whether it supported testing. However, this pattern cannot be cleanly attributed to a single component, since the non-agent and agent conditions differ on multiple dimensions simultaneously, such as task framing, attentional load, output modality and browser interaction. Second, relaxing the maximum step limit substantially increases task completion rates, while also leading the agent to consume more steps to complete the same tasks under a looser budget. Cost-effectiveness should therefore also be considered in design. For an example task, the resulting computing cost was approximately $1.72 for Claude Opus 4.5 (30 steps), $0.65 for Gemini 3 Pro (40 steps), and $0.17 for Gemini 3 Flash (30 steps); or roughly $0.057, $0.016, and $0.006 per step, respectively, after accounting for prompt caching. Even though the lighter model needed a larger max step limit to reach completion rates comparable to the frontier models, its overall cost could remain lower. It should be noted that these estimates reflect only the computing cost of individual agent runs in our sandbox and should not be taken to approximate the cost of real-world deployment, which must also account for operational and maintenance costs and the labor cost of human supervision. Characterizing these cost components will be an important consideration for future work.

Third, while augmenting agent prompts with error-specific information improved performance on the targeted error types, the targeted prompt operates narrowly on its intended category rather than conferring general improvements in agent reasoning. Prior work shows that prompt engineering is highly context-and task-dependent, often improving some subtasks while degrading others, and remains sensitive to small variations in phrasing or format.^24^ This brittleness makes exhaustive prompt engineering an impractical strategy for clinical deployment, as it would require anticipating every possible failure mode in advance.

In practice, a structured onboarding or “shadowing” phase, during which the agent observes real submission workflows, encounters errors, and iteratively refines its own prompts or skill documents, is likely necessary to achieve broad coverage.^25,26^ Finally, our results suggest that reliable error detection and form submission may be fundamentally difficult to achieve within a single autonomous agent at current model capability. A controlled, modular workflow may be a better architecture for clinical deployment, given that PA submission for ES/GS itself is a coordinated team effort: administrative staff compile patient information from the EHR, a genetic counselor or PA specialist reviews documentation for completeness and clinical appropriateness, and a designated submitter enters the finalized form.^27,28^ A multi-agent pipeline would mirror this workflow naturally, and emerging evidence suggests that multi-agent designs outperform monolithic LLM agents in clinical tasks.^29,30^ This staged design can also position a human reviewer as a deliberate participant rather than a final rubber stamp: the agent’s output should first be presented to a geneticist in a human-readable format, with each identified issue paired to its supporting evidence, before any submission is triggered.

However, whether such a pipeline delivers net time savings in practice will depend on how human oversight is designed, including at which points the agent pauses for review, when a human intervenes or halts the trajectory, and how much of the output must be verified. These are open human-AI interaction questions that warrant dedicated future work.

Several limitations bound the interpretation of our findings. First, the EHR environment was deliberately simplified and understates the burden of a production EHR: the information required for a single payer’s PA form was consolidated into one structured record, whereas in practice some of these fields would not appear directly in clinical documentation. In addition, some generated fields carry artifacts of the synthetic data generation process that do not conform to real-world conventions (e.g., randomly generated NPI numbers and addresses) that introduce noise and make it harder to distinguish genuine reasoning failures from responses to unrealistic data. The use of synthetic data itself, however, was a deliberate design choice rather than solely a constraint: exposing an unvalidated agentic workflow to real patient records would carry privacy and safety risks, and synthetic profiles allow error types to be injected and controlled systematically. Building on our findings, a clinically validated cohort, for example, one drawn from de-identified real PA submissions with clinical reviews to define compliant and deficient cases, will be an important direction for future work. Second, we evaluated agents under a single baseline prompt template and did not systematically test alternative prompting strategies which could meaningfully shift the behaviors we observed. In addition, the agent and non-agent prompts in Ablation 2 were not identically worded because of the difference in setting. The agent prompt instructed the agent to “verify all required fields and then directly submit,” framing submission as the default action with halting as the exception, whereas the non-agent prompt asked the model to “determine if the pre-authorization form should be submitted,” framing the task as an open decision. Since this difference in framing could bias the agent toward submission, the contrast between the two settings may also partly reflect prompt wording rather than agentic scaffolding itself. Finally, the models evaluated in this study only represent the agent capabilities available at the time of the study. Given the pace of model development, the specific rates we report may not hold for newer models, and the sandbox is intended to support re-evaluation as new models are released rather than serve as a fixed ranking.

## Declaration

### Data Availability

The source code of the sandbox and experiment runs is available at https://github.com/stormliucong/wes-wgs-pa-app.

## Author Contributions

Conceptualization: C.L., M.N.; Data curation: M.N.; Formal analysis: M.N., C.L.; Funding acquisition: C.L.; Investigation: M.N., C.L.; Methodology: C.L., M.N.; Project administration: M.N., C.L.; Resources: C.L.; Software: C.L., M.N., P.A.; Supervision: C.L.; Validation: C.L., P.A., J.W., M.L.; Visualization: M.N., C.L., C.W., K.W.; Writing-original draft: M.N., C.L.; Writing-review & editing: C.L., W.C., M.L., J.W., C.W., R.L., K.W., P.A.

## Ethics Declaration

Not applicable, there is no human subject or real patient data involved in the study.

## Declaration of generative AI and AI-assisted technologies in the manuscript preparation process

During the preparation of this work the author(s) used Claude and ChatGPT in order to improve phrasing and the flow of writing. After using this tool/service, the author(s) reviewed and edited the content as needed and take(s) full responsibility for the content of the publication.

## Supporting information

Prompt library, Figure S1, Figure S2, Table S1, Table S2, Table S3, Table S4

## References

1. Marcus BS, Bansal N, Saef J, et al. Burden with No Benefit: Prior Authorization in Congenital Cardiology. Pediatr Cardiol. 2024;45(1):100–106. doi:10.1007/s00246-023-03255-1

2. Sahni NR, Istvan B, Stafford C, Cutler D. Perceptions of prior authorization burden and solutions. Health Affairs Scholar. 2024;2(9):qxae096. doi:10.1093/haschl/qxae096

3. Some Medicare Advantage Organization Denials of Prior Authorization Requests Raise Concerns About Beneficiary Access to Medically Necessary Care. Office of Inspector General | Government Oversight | U.S. Department of Health and Human Services. April 27, 2022. Accessed June 1, 2026. https://oig.hhs.gov/reports/all/2022/some-medicare-advantage-organization-denials-of-prior-authorization-requests-raise-concerns-about-beneficiary-access-to-medically-necessary-care/

4. kffmichellel. Claims Denials and Appeals in ACA Marketplace Plans in 2024. KFF. March 24, 2026. Accessed April 29, 2026. https://www.kff.org/patient-consumer-protections/claims-denials-and-appeals-in-aca-marketplace-plans-in-2024/

5. Kim MK, Rouphael C, McMichael J, Welch N, Dasarathy S. Challenges in and Opportunities for Electronic Health Record-Based Data Analysis and Interpretation. Gut and Liver. 2024;18(2):201–208. doi:10.5009/gnl230272

6. Fixing prior auth: Nearly 40 prior authorizations a week is way too many. American Medical Association. April 24, 2025. Accessed April 29, 2026. https://www.ama-assn.org/practice-management/prior-authorization/fixing-prior-auth-nearly-40-prior-authorizations-week-way

7. Stark Z, Schofield D, Alam K, et al. Prospective comparison of the cost-effectiveness of clinical whole-exome sequencing with that of usual care overwhelmingly supports early use and reimbursement. Genet Med. 2017;19(8):867–874. doi:10.1038/gim.2016.221

8. Clark MM, Stark Z, Farnaes L, et al. Meta-analysis of the diagnostic and clinical utility of genome and exome sequencing and chromosomal microarray in children with suspected genetic diseases. NPJ Genom Med. 2018;3:16. doi:10.1038/s41525-018-0053-8

9. Smith HS, Franciskovich R, Lewis AM, et al. Outcomes of prior authorization requests for genetic testing in outpatient pediatric genetics clinics. Genet Med. 2021;23(5):950–955. doi:10.1038/s41436-020-01081-x

10. Zion TN, Berrios CD, Cohen ASA, et al. Insurance denials and diagnostic rates in a pediatric genomic research cohort. Genet Med. 2023;25(5):100020. doi:10.1016/j.gim.2023.100020

11. Anderson KE, Darden M, Jain A. Improving Prior Authorization in Medicare Advantage. JAMA. 2022;328(15):1497–1498. doi:10.1001/jama.2022.17732

12. From Transactions to Trust: Building Better Care Through Healthcare Automation. 2025. Accessed October 16, 2026. https://www.caqh.org/hubfs/Index/2024%20Index%20Report/CAQH_IndexReport_2024_FINAL.pdf

13. Yao S, Zhao J, Yu D, et al. ReAct: Synergizing Reasoning and Acting in Language Models. arXiv. Preprint posted online March 10, 2023:arXiv:2210.03629. doi:10.48550/arXiv.2210.03629

14. He H, Yao W, Ma K, et al. WebVoyager: Building an End-to-End Web Agent with Large Multimodal Models. arXiv. Preprint posted online June 6, 2024:arXiv:2401.13919. doi:10.48550/arXiv.2401.13919

15. Zhou S, Xu FF, Zhu H, et al. WebArena: A Realistic Web Environment for Building Autonomous Agents. arXiv. Preprint posted online April 16, 2024:arXiv:2307.13854. doi:10.48550/arXiv.2307.13854

16. Jiang Y, Black KC, Geng G, et al. MedAgentBench: A Virtual EHR Environment to Benchmark Medical LLM Agents. NEJM AI. 2025;2(9). doi:10.1056/AIdbp2500144

17. Bedi S, Welch R, Steinberg E, et al. HealthAdminBench: Evaluating Computer-Use Agents on Healthcare Administration Tasks. arXiv.org. April 10, 2026. Accessed October 16, 2026. https://arxiv.org/abs/2604.09937v1

18. Aronson SJ, Machini K, Shin J, et al. GPT-4 Performance, Nondeterminism, and Drift in Genetic Literature Review. NEJM AI. 2024;1(9):AIcs2400245. doi:10.1056/AIcs2400245

19. Bowman S. Impact of electronic health record systems on information integrity: quality and safety implications. Perspect Health Inf Manag. 2013;10(Fall):1c.

20. Bell SK, Delbanco T, Elmore JG, et al. Frequency and Types of Patient-Reported Errors in Electronic Health Record Ambulatory Care Notes. JAMA Netw Open. 2020;3(6):e205867. doi:10.1001/jamanetworkopen.2020.5867

21. Drouin A, Gasse M, Caccia M, et al. WorkArena: how capable are web agents at solving common knowledge work tasks? In: Proceedings of the 41st International Conference on Machine Learning. Vol 235. ICML’24. JMLR.org; 2024:11642-11662.

22. Levy I, Wiesel B, Marreed S, Oved A, Yaeli A, Shlomov S. ST-WebAgentBench: A Benchmark for Evaluating Safety and Trustworthiness in Web Agents. arXiv. Preprint posted online March 2, 2026:arXiv:2410.06703. doi:10.48550/arXiv.2410.06703

23. Kirichenko P, Ibrahim M, Chaudhuri K, Bell S. AbstentionBench: Reasoning LLMs Fail on Unanswerable Questions. In: 2025. Accessed April 30, 2026. https://openreview.net/forum?id=OkHC30LLpO&referrer=%5Bthe%20profile%20of%20Samuel%20J.%20Bell%5D(%2Fprofile%3Fid%3D~Samuel_J_Bell1)

24. Chai M, Zomorrodi AR. Prompt engineering does not universally improve Large Language Model performance across clinical decision-making tasks. arXiv. Preprint posted online December 28, 2025:arXiv:2512.22966. doi:10.48550/arXiv.2512.22966

25. Madaan A, Tandon N, Gupta P, et al. Self-Refine: Iterative Refinement with Self-Feedback. arXiv. Preprint posted online May 25, 2023:arXiv:2303.17651. doi:10.48550/arXiv.2303.17651

26. Mokssit Y, Ravi K, Nie M, Kim J, Liu C. FHIR-AgentEval: a modular sandbox for benchmarking clinical LLM agents with memory-augmented configurations. *npj Digit Med*. Published online October 10, 2026. doi:10.1038/s41746-026-03348-0

27. Uhlmann WR, Schwalm K, Raymond VM. Development of a Streamlined Work Flow for Handling Patients’ Genetic Testing Insurance Authorizations. Journal of Genetic Counseling. 2017;26(4):657–668. doi:10.1007/s10897-017-0098-3

28. Bajguz D, Danylchuk NR, Czarniecki M, Selig JP, Sutphen R, Kaylor J. Utilization of genetic testing: Analysis of 4,499 prior authorization requests for molecular genetic tests at four US regional health plans. Journal of Genetic Counseling. 2022;31(3):771–780. doi:10.1002/jgc4.1543

29. Chen X, Yi H, You M, et al. Enhancing diagnostic capability with multi-agents conversational large language models. npj Digit Med. 2025;8(1):159. doi:10.1038/s41746-025-01550-0

30. Klang E, Omar M, Raut G, et al. Orchestrated multi agents sustain accuracy under clinical-scale workloads compared to a single agent. npj Health Syst. 2026;3(1):23. doi:10.1038/s44401-026-00077-0

